# Machine Learning–based Prediction of Pacing-Induced Cardiomyopathy in Patients with Atrioventricular Block Underlying Heart Failure with Mildly Reduced or Preserved Ejection Fraction

**DOI:** 10.64898/2026.10.09.26365179

**Authors:** Jang-Won Lee, Ungjeong Do, Hyue Mee Kim, Min Jea Choi, Seung Yong Shin, Seung Park, Ki-Woon Kang

**Author notes:** **Corresponding Authors:** Ki-Woon Kang, MD, PhD, Division of Cardiology, Chung-Ang University Hospital, College of Medicine, Chung-Ang University, 102 Heukseok-ro, Dongjak-gu, Seoul 06973, Republic of Korea. and MitoKey Co., Ltd., 84 Heukseok-ro,Dongjak-gu, Seoul, 06974, South Korea; Seung Park, PhD, Graduate School of Public Health and Healthcare Management, The Catholic University of Korea, Seoul, Republic of Korea.

## Abstract

**Background:** Pacing-induced cardiomyopathy (PICM) is a major complication of chronic right ventricular pacing and develops in up to 25% of patients. Reliable prediction of PICM may facilitate individualized pacing strategy selection; however, currently available approaches demonstrate limited discriminative performance. This study aimed to develop and validate machine learning (ML) models for predicting PICM in patients with atrioventricular block (AVB) underlying heart failure with mildly reduced or preserved ejection fraction.

**Methods:** In this multicenter prospective study derived from the RISK-PICM registry (January 2002 to January 2026), 125 patients with AVB who underwent pacemaker implantation were analyzed. PICM was defined as a ≥10% reduction in left ventricular ejection fraction (LVEF) resulting in an LVEF <50%. Four ML algorithms, including logistic regression, support vector machine, random forest, and XGBoost, were trained using stratified 4-fold cross-validation with hyperparameter optimization and model discrimination was evaluated.

**Results:** PICM developed in 41 patients (32.8%) during a median follow-up of 4.8 years. Male sex, pre-implant minimum QTc interval, and post-implant maximum QRS duration were significantly associated with PICM in univariable Cox regression. XGBoost achieved the highest discriminative performance (AUROC 0.831; sensitivity 0.805; specificity 0.845). Feature importance analysis identified baseline LVEF and post-implant maximum QRS duration as the most influential features. Model-derived risk stratification significantly differentiated high-risk and low-risk groups on Kaplan-Meier analysis (*P* < 0.001).

**Conclusions:** ML-based models, particularly XGBoost, demonstrated clinically meaningful performance for PICM and may support individualized pacing strategy selection in patients with AVB and underlying heart failure with mildly reduced or preserved ejection fraction.

**Clinical Perspective:** *What Is New:*

- In pacing-dependent AVB patients with HFmrEF or HFpEF, ensemble ML (XGBoost) predicted PICM with an AUROC of 0.831.
- Model-derived risk stratification separated PICM-free survival (log-rank P<0.001).

*What Are the Clinical Implications:*

- An ML model combining baseline LVEF, paced QRS duration, and the full parameter set may guide individualized choice between right ventricular and conduction system pacing in AVB patients with HFmrEF or HFpEF.
- Using paced QRS duration combined with baseline LVEF during temporary pacing in AVB patients with HFmrEF or HFpEF, this risk estimate could inform pacing-strategy selection before permanent implantation, enabling pre-emptive use of conduction system pacing in high-risk patients.

## Introduction

Chronic right ventricular pacing (RVP) remains the established standard therapy for symptomatic bradycardia requiring pacing rhythm ^1,2^. Contemporary expert consensus statements endorse remote monitoring as the standard follow-up strategy for patients with permanent pacemakers, with in-person evaluations often performed as infrequently as once every one to two years. However, this consensus paradigm relies primarily on device-transmitted alerts and does not routinely incorporate the electrocardiography (ECG) and left ventricular ejection fraction (LVEF) assessment necessary for the early detection of pacing-induced cardiomyopathy (PICM). Importantly, PICM develops in approximately 12%–25% of patients undergoing chronic RVP and may progress silently until overt atrial fibrillation (AF) and heart failure (HF) become clinically apparent, particularly in patients with atrioventricular block (AVB) ^2–5^.

To reduce the risk of PICM, conduction system pacing (CSP) has emerged as a physiologically favorable alternative that preserves native ventricular activation and maintains electrical and mechanical synchrony ^6–8^. Although CSP is increasingly preferred in patients with HF with reduced EF (HFrEF), in whom chronic RVP is now generally discouraged, evidence supporting CSP in patients with HF with preserved EF (HFpEF) or HF with mildly reduced EF (HFmrEF) remains limited because of the relative paucity of prospective data ^9,10^.

Recent consensus statements suggest that CSP may be considered in patients with HFpEF or HFmrEF who are expected to require a ventricular pacing burden >20% ^11,12^. Nevertheless, compared with conventional biventricular pacing (BiVP) or RVP, CSP remains technically more demanding, requiring longer procedural times, a steeper learning curve, and more frequent lead revisions in the pacing dependent rhythm ^13,14^. In contrast, conventional RVP is associated with greater procedural simplicity, shorter implantation times, and more favorable lead profiles, thereby remaining a clinically practical and cost-effective pacing strategy, particularly in patients at relatively low risk for PICM. Consequently, accurate risk stratification for PICM is critical to support individualized selection between conventional RVP and CSP in the pacing dependent rhythm. Previous studies have identified several factors associated with PICM development, including male sex, lower pre-implant LVEF, prolonged QRS duration (QRSd), and higher RVP burden ^15–17^. However, these variables individually demonstrate only modest predictive strength and insufficient discriminatory capacity when considered in isolation. To date, no validated multivariable prediction model integrating these clinical and electrocardiographic parameters has been established for reliable estimation of PICM risk.

To address the limited predictive performance of conventional statistical approaches, a prior study applied convolutional neural networks (CNN) for PICM prediction; however, the model demonstrated relatively low sensitivity and was intrinsically limited by the tabular structure of the clinical and electrocardiographic data ^18^. Accordingly, the present study aimed to develop and validate machine learning (ML)-based prediction models for PICM in AVB patients with HFpEF or HFmrEF. We hypothesized that ensemble-based ML models would achieve superior discriminative performance for identifying patients at high risk for PICM and would provide a realistic estimate of achievable discrimination and thereby facilitate individualized pacing strategy selection.

## Methods

### Study Design and Population

This prospective registry is a multicenter cohort study conducted at three tertiary referral centers in South Korea, including Eulji University Hospital, Chungbuk National University Hospital, and Chung-Ang University Hospital. The study protocol was approved by the institutional review board at each participating institution (approval number 2018-006-474), and all participants provided written informed consent. A total of 697 consecutive patients who underwent permanent pacemaker implantation were enrolled, with prospective enrollment since January 2002; the registry was formally named RISK-PICM and registered at ClinicalTrials.gov (NCT03475498) in 2018. Enrollment for the present analysis continued through January 2026. 125 patients with complete pre- and post-implant 12-lead ECGs and echocardiographic datasets were included in the present analysis. Patients were eligible for inclusion if they had a confirmed diagnosis of AVB requiring chronic RVP. Exclusion criteria included; pre-implant HFrEF (LVEF ≤40%); pre-existing persistent atrial fibrillation; myocardial infarction at the time of implantation; ventricular pacing burden <80% to ensure pacing dependency; and tachycardia-bradycardia syndrome in Supplemental Figure.

Clinical data were entered into a standardized web-based electronic database incorporating built-in validation procedures for key variables. Data management was performed using the internet-based Clinical Research and Trial management system (iCReaT; http://icreat.nih.go.kr), a government-supported clinical research platform.

### Electrocardiographic and Echocardiographic Assessment

Pre-implant ECG parameters were obtained from 12-lead ECGs recorded during confirmed AVB within one year before pacemaker implantation. For each patient, at least three separate ECG recordings were reviewed to determine the mode (most frequently observed), maximum, and minimum values for QRSd and corrected QT interval (QTc). Post-implant ECG parameters were derived from ECGs demonstrating stable ventricular paced rhythm, and at least three separate recordings were similarly selected to determine the corresponding mode, maximum, and minimum ECG measurements.

Pre-implant echocardiographic parameters were obtained from the transthoracic echocardiogram (TTE) performed closest to the implantation date within one year before the procedure. For post-implant assessment, the TTE demonstrating a clinically meaningful decline in LVEF was selected; if no substantial reduction was observed, the most recent follow-up TTE was analyzed. Echocardiographic studies were excluded if the reduction in LVEF was attributable to an alternative identifiable cause, including acute ischemic heart disease or progressive valvular disease.

### Definition of Heart Failure Categorization and Pacing-Induced Cardiomyopathy

HF was defined as a clinical syndrome characterized by symptoms and/or signs resulting from structural or functional cardiac abnormalities, supported by elevated natriuretic peptide levels or objective evidence of pulmonary or systemic congestion. Patients were further categorized according to LVEF into three HF subtypes in accordance with current guidelines ^19,20^: HFrEF (LVEF ≤40%), HFmrEF (LVEF 41%–49%), and HFpEF (LVEF ≥50%). In the absence of a universally accepted definition, PICM was defined as a reduction in LVEF of at least 10% from baseline, resulting in a post-implant LVEF <50%, consistent with the most commonly adopted definition in prior studies ^17,21^. Time-to-event was defined as the interval from pacemaker implantation to documented PICM occurrence or the last clinical follow-up, whichever occurred first.

### Data Preprocessing

To minimize the risk of data leakage, all preprocessing procedures, including imputation and standardization, were incorporated within the cross-validation pipeline to preserve analytical consistency and reproducibility. The primary outcome was PICM occurrence, treated as a binary variable, whereas time-to-event was defined as the duration of clinical follow-up in months. Records with missing data for either of these core variables were excluded before analysis.

Missing values among the remaining variables were handled according to variable type. Binary categorical variables, including sex, hypertension, diabetes mellitus, and alcohol consumption, were imputed using the mode. Continuous variables, including pre- and post-implant ECG parameters, hemoglobin concentration, and estimated glomerular filtration rate, were imputed using mean values under the assumption of approximate normal distribution.

Imputation parameters (e.g., mean and mode) and standardization parameters (e.g., mean and standard deviation) were derived exclusively from the training folds and subsequently applied to the validation folds during each cross-validation iteration. Following imputation, categorical variables were converted into numerical representations using one-hot encoding, with one reference category removed to minimize multicollinearity. Continuous variables were standardized using z-score normalization to prevent differences in measurement scale from disproportionately influencing model training.

### Statistical Analysis

Baseline characteristics were compared between the PICM and non-PICM groups. Continuous variables were presented as mean ± standard deviation and compared using independent-samples t-tests. Categorical variables were expressed as frequencies with % and compared using the chi-square test or Fisher’s exact test, as appropriate. Cox proportional hazards regression analysis was performed to identify variables associated with PICM occurrence, and the results were reported as hazard ratios with 95% confidence intervals. The proportional hazards assumption was assessed using Schoenfeld residuals. A two-sided P value <0.05 was considered statistically significant.

### Machine Learning Model Development

To develop predictive models for PICM occurrence, stratified four-fold cross-validation was implemented to preserve the distribution of events and non-events across training and validation datasets and thereby minimize sampling bias. Four supervised ML algorithms were evaluated, including logistic regression, support vector machine (SVM), random forest, and extreme gradient boosting (XGBoost).

Within each fold, hyperparameter optimization was performed using Optuna, a Bayesian optimization framework designed to efficiently explore high-dimensional parameter spaces. The optimization objective was to maximize the area under the receiver operating characteristic curve (AUROC) using an internal validation subset derived from the training data while maintaining complete separation of the held-out validation fold during the tuning process. All models were independently trained and evaluated within each outer cross-validation fold, and fold-specific performance metrics were subsequently averaged to generate robust estimates of model generalizability.

To improve model interpretability and reduce the risk of overfitting, feature selection was performed on the basis of statistical significance in univariate analyses, with only variables demonstrating a *P* value <0.05 in between-group comparisons included in model development. Model discrimination was evaluated using receiver operating characteristic (ROC) curve analysis, and the optimal classification threshold within each fold was determined by maximizing Youden’s J statistic. Sensitivity, specificity, and AUROC values were calculated at the corresponding threshold, and mean performance metrics across all folds were reported. Feature importance analyses were additionally performed for the random forest and XGBoost models to identify variables contributing most substantially to predictive performance.

### Risk Stratification and Survival Analysis

Predicted risk probabilities generated by the best-performing model were used to stratify patients into high-risk and low-risk groups for PICM development. The optimal threshold for risk stratification was determined by maximizing Youden’s J statistic within each fold. Kaplan– Meier survival analyses were subsequently performed for the two risk groups using time-to-event data defined by follow-up duration in months, and differences in PICM-free survival were compared using the log-rank test.

## Results

### Baseline Characteristics

Among the 697 patients enrolled in the registry, 125 satisfied all eligibility criteria and had complete datasets available for the final analysis. During a median follow-up duration of 4.8 years (interquartile range, 2.1–8.6 years), PICM developed in 41 patients (32.8%), whereas 84 patients (67.2%) did not develop PICM.

Baseline demographic and clinical characteristics stratified according to PICM status are summarized in Table 1. Mean age did not significantly differ between the PICM and non-PICM groups (75.6 ± 11.6 years vs. 74.3 ± 11.6 years; *P* = 0.563). However, proportion of male were significantly more prevalent in the PICM group than in the non-PICM group (56.1% vs. 32.5%; *P* = 0.012).

**Table 1.** Baseline characteristics of patients with and without pacing-induced cardiomyopathy.

|  | Overall<br>n=125 | Without PICM<br>n=84 | With PICM<br>n=41 | P value |
| --- | --- | --- | --- | --- |
| Age, years | 74.8 ± 11.6 | 74.3 ± 11.6 | 75.6 ± 11.6 | 0.563 |
| Male, n (%) | 50 (40.0) | 27 (32.1) | 23 (56.1) | 0.012* |
| Hypertension, n (%) | 74 (59.2) | 48 (57.1) | 26 (63.4) | 0.595 |
| Diabetes mellitus, n (%) | 29 (23.2) | 22 (26.2) | 7 (17.1) | 0.228 |
| Coronary artery disease, n (%) | 8 (6.4) | 5 (6.0) | 3 (7.3) | 1.000 |
| End stage renal disease, n (%) | 3 (2.5) | 2 (2.5) | 1 (2.5) | 1.000 |
| Ischemic heart disease, n (%) | 5 (4.2) | 4 (5.0) | 1 (2.5) | 0.664 |
| Stroke, n (%) | 6 (5.0) | 3 (3.8) | 3 (7.5) | 0.399 |
| Alcohol, n (%) | 17 (14.2) | 9 (11.2) | 8 (20.0) | 0.195 |
| Smoking, n (%) | 11 (9.2) | 8 (10.0) | 3 (7.5) | 0.750 |
| Pre-implant hemoglobin, g/dL | 12.5 ± 2.0 | 12.4 ± 2.1 | 12.5 ± 1.7 | 0.741 |
| Pre-implant GFR (mean ± SD), | 81.6 ± 29.3 | 80.9 ± 27.3 | 82.9 ± 33.0 | 0.744 |
| Pre-implant mode QRSd, ms | 112.1 ± 27.1 | 109.7 ± 27.7 | 117.0 ± 25.6 | 0.144 |
| Pre-implant QTc, ms | 448.5 ± 51.1 | 448.8 ± 54.5 | 447.9 ± 43.9 | 0.922 |
| Pre-implant max QRSd, ms | 122.6 ± 30.9 | 120.1 ± 31.0 | 127.7 ± 30.5 | 0.197 |
| Pre-implant max QTc, ms | 462.9 ± 55.9 | 464.4 ± 56.4 | 459.6 ± 55.2 | 0.650 |
| Pre-implant min QRSd, ms | 105.3 ± 26.0 | 104.1 ± 26.8 | 107.9 ± 24.5 | 0.427 |
| Pre-implant min QTc, ms | 438.1 ± 51.7 | 446.3 ± 52.6 | 421.3 ± 46.0 | 0.008* |
| Post-implant mode QRSd, ms | 150.8 ± 25.9 | 147.8 ± 27.4 | 157.0 ± 21.5 | 0.045* |
| Post-implant mode QTc, ms | 488.9 ± 42.9 | 487.0 ± 45.5 | 492.8 ± 37.5 | 0.453 |
| Post-implant max QRSd, ms | 168.0 ± 28.9 | 162.0 ± 31.5 | 180.3 ± 17.6 | <0.001* |
| Post-implant max QTc, ms | 511.8 ± 47.6 | 507.3 ± 52.1 | 521.0 ± 35.4 | 0.086 |
| Post-implant min QRSd, ms | 129.9 ± 28.1 | 127.6 ± 29.5 | 134.7 ± 24.8 | 0.157 |
| Post-implant min QTc, ms | 445.6 ± 65.9 | 446.6 ± 58.8 | 443.5 ± 79.4 | 0.821 |
| Pre-implant EF, % | 65.0 ± 9.2 | 64.7 ± 10.0 | 65.8 ± 7.4 | 0.527 |
| Post-implant EF, % | 55.1 ± 11.4 | 60.9 ± 7.1 | 43.1 ± 9.0 | <0.001* |

\* Statistically significant. Continuous variables are presented as mean ± SD.
PICM: pacing-induced cardiomyopathy; Mode: most frequently observed value among ECG parameters; QRSd: QRS duration; QTc: corrected QT interval; EF: ejection fraction.

The prevalence of comorbid conditions, including hypertension (63.4% vs. 57.1%; *P* = 0.595), diabetes mellitus (17.1% vs. 26.2%; *P* = 0.228), coronary artery disease (7.3% vs. 6.0%; *P* = 1.000), and ischemic heart disease (2.5% vs. 5.0%; *P* = 0.664), was comparable between groups. Similarly, laboratory parameters associated with HF progression, including hemoglobin concentration (12.5 ± 1.7g/dL vs. 12.4 ± 2.1g/dL; *P* = 0.741) and estimated glomerular filtration rate (82.9 ± 33.0mL/min/1.73 m² vs. 80.9 ± 27.3mL/min/1.73 m²; *P* = 0.744), did not significantly differ between the groups.

### Electrocardiographic and Echocardiographic Data

Pre-implant and post-implant ECG parameters are summarized in Table 1. Among the pre-implant ECG variables, the minimum QTc interval was significantly shorter in the PICM group than in the non-PICM group (421.3 ± 46.0ms vs. 446.3 ± 52.6ms; *P* = 0.008), whereas no significant between-group differences were observed for the remaining pre-implant QRSd and QTc parameters.

More pronounced differences were observed in post-implant ECG parameters. The mode paced QRSd was significantly longer in the PICM group than in the non-PICM group (157.0 ± 21.5ms vs. 147.8 ± 27.4ms; *P* = 0.045), with the greatest difference observed for the maximum paced QRSd (180.3 ± 17.6ms vs. 162.0 ± 31.5ms; *P* < 0.001). Baseline pre-implant LVEF was comparable between the non-PICM and PICM groups (64.7 ± 10.0% vs. 65.8 ± 7.4%; *P* = 0.527). In contrast, post-implant LVEF was significantly lower in the PICM group than in the non-PICM group (43.1 ± 9.0% vs. 60.9 ± 7.1%; *P* < 0.001).

### Cox Proportional Hazards Analysis

Univariable Cox proportional hazards regression analysis was performed to identify clinical and electrocardiographic variables associated with PICM occurrence (Table 2). Three variables demonstrated statistically significant associations with PICM development. Male sex was associated with an approximately twofold higher risk of PICM occurrence (HR 2.01; 95% CI, 1.08–3.72; *P* = 0.027). The pre-implant minimum QTc interval demonstrated an inverse association with PICM risk (HR 0.99 per 1-ms increase; 95% CI, 0.98–1.00; *P* = 0.016). Among the ECG variables, post-implant maximum QRSd emerged as the strongest predictor (HR 1.02 per 1-ms increase; 95% CI, 1.01–1.04; *P* = 0.001).

**Table 2.** Cox proportional hazards regression analysis for pacing-induced cardiomyopathy occurrence.

| Variable | HR | 95% CI | P value |
| --- | --- | --- | --- |
| Age, per year | 1.02 | 0.98 - 1.05 | 0.330 |
| Sex, male | 2.01 | 1.08 - 3.72 | 0.027* |
| Hypertension | 1.24 | 0.64 - 2.37 | 0.525 |
| Diabetes mellitus | 0.70 | 0.31 - 1.57 | 0.383 |
| Coronary artery disease | 1.82 | 0.56 - 5.96 | 0.322 |
| End stage renal disease | 1.55 | 0.21 - 11.40 | 0.667 |
| Ischemic heart disease | 0.66 | 0.09 - 4.84 | 0.686 |
| Stroke | 1.89 | 0.58 - 6.16 | 0.288 |
| Alcohol | 1.75 | 0.80 - 3.79 | 0.159 |
| Smoking | 0.68 | 0.21 - 2.22 | 0.528 |
| Pre-implant hemoglobin, g/L | 1.00 | 0.86 - 1.16 | 0.976 |
| Pre-implant GFR | 1.00 | 0.99 - 1.01 | 0.958 |
| Pre-implant mode QRSd, ms | 1.01 | 0.99 - 1.02 | 0.369 |
| Pre-implant mode QTc, ms | 1.00 | 0.99 - 1.01 | 0.922 |
| Pre-implant max QRSd, ms | 1.01 | 1.00 - 1.02 | 0.274 |
| Pre-implant max QTc, ms | 1.00 | 0.99 - 1.01 | 0.923 |
| Pre-implant min QRSd, ms | 1.00 | 0.99 - 1.01 | 0.686 |
| Pre-implant min QTc, ms | 0.99 | 0.98 - 1.00 | 0.016* |
| Post-implant mode QRSd, ms | 1.01 | 1.00 - 1.02 | 0.134 |
| Post-implant mode QTc, ms | 1.00 | 0.99 - 1.01 | 0.682 |
| Post-implant max QRSd, ms | 1.02 | 1.01 - 1.04 | 0.001* |
| Post-implant max QTc, ms | 1.01 | 1.00 - 1.01 | 0.096 |
| Post-implant min QRSd, ms | 1.01 | 0.99 - 1.02 | 0.367 |
| Post-implant min QTc, ms | 1.00 | 0.99 - 1.00 | 0.522 |

\* Statistically significant. QRSd: QRS duration; QTc: corrected QT interval.

Other baseline clinical characteristics, including age (HR 1.02; *P* = 0.330), hypertension (HR 1.24; *P* = 0.525), diabetes mellitus (HR 0.70; *P* = 0.383), hemoglobin concentration, and estimated glomerular filtration rate, were not significantly associated with PICM occurrence in this cohort.

### Machine Learning Model Performance

The discriminative performance of the four ML models evaluated using stratified four-fold cross-validation is summarized in Table 3. Among the evaluated algorithms, XGBoost demonstrated the highest overall discriminative performance, achieving a mean AUROC of 0.831 with balanced sensitivity (0.805) and specificity (0.845). Random forest demonstrated comparable performance, with an AUROC of 0.806 and the highest specificity among all models (0.857), while maintaining acceptable sensitivity (0.683).

**Table 3.** Comparison of machine learning model performance.

| Model | AUROC | Sensitivity | Specificity |
| --- | --- | --- | --- |
| Logistic Regression | 0.732 | 0.585 | 0.786 |
| SVM | 0.740 | 0.732 | 0.679 |
| Random Forest | 0.806 | 0.683 | 0.857 |
| XGBoost | 0.831 | 0.805 | 0.845 |

SVM achieved an AUROC of 0.740 with a sensitivity of 0.732 and specificity of 0.679, indicating moderate but comparatively less balanced predictive performance. Logistic regression demonstrated the lowest overall discriminative ability, with an AUROC of 0.732, sensitivity of 0.585, and specificity of 0.786. ROC curves for all four models are presented in Figure 1.

**Figure 1.**
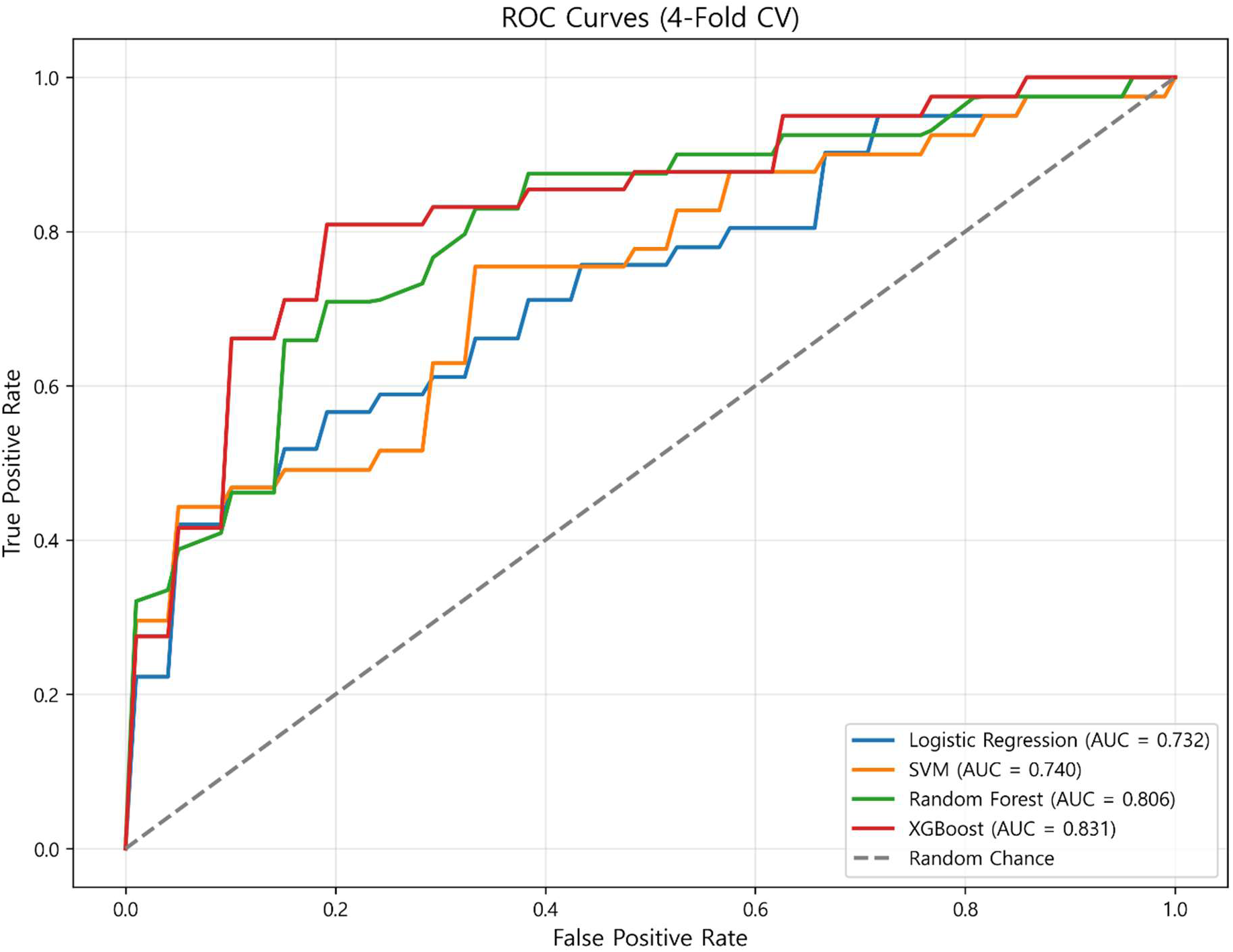
Receiver operating characteristic (ROC) curves comparing the predictive performance of machine learning models for pacing-induced cardiomyopathy. Four models were evaluated using stratified 4-fold cross-validation. XGBoost demonstrated the highest discriminative performance (AUROC = 0.831), followed by random forest (AUROC = 0.806), support vector machine (SVM; AUROC = 0.740), and logistic regression (AUROC = 0.732). The dashed diagonal line represents random classification performance (AUROC = 0.50). AUROC = area under the receiver operating characteristic curve; SVM = support vector machine.

### Feature Importance Analysis

Feature importance rankings derived from the random forest and XGBoost models demonstrated substantial concordance, as illustrated in Figure 2. Baseline LVEF emerged as the most influential predictor of PICM in both models, accounting for 23.5% of total feature importance in the random forest model and 28.3% in the XGBoost model.

**Figure 2.**
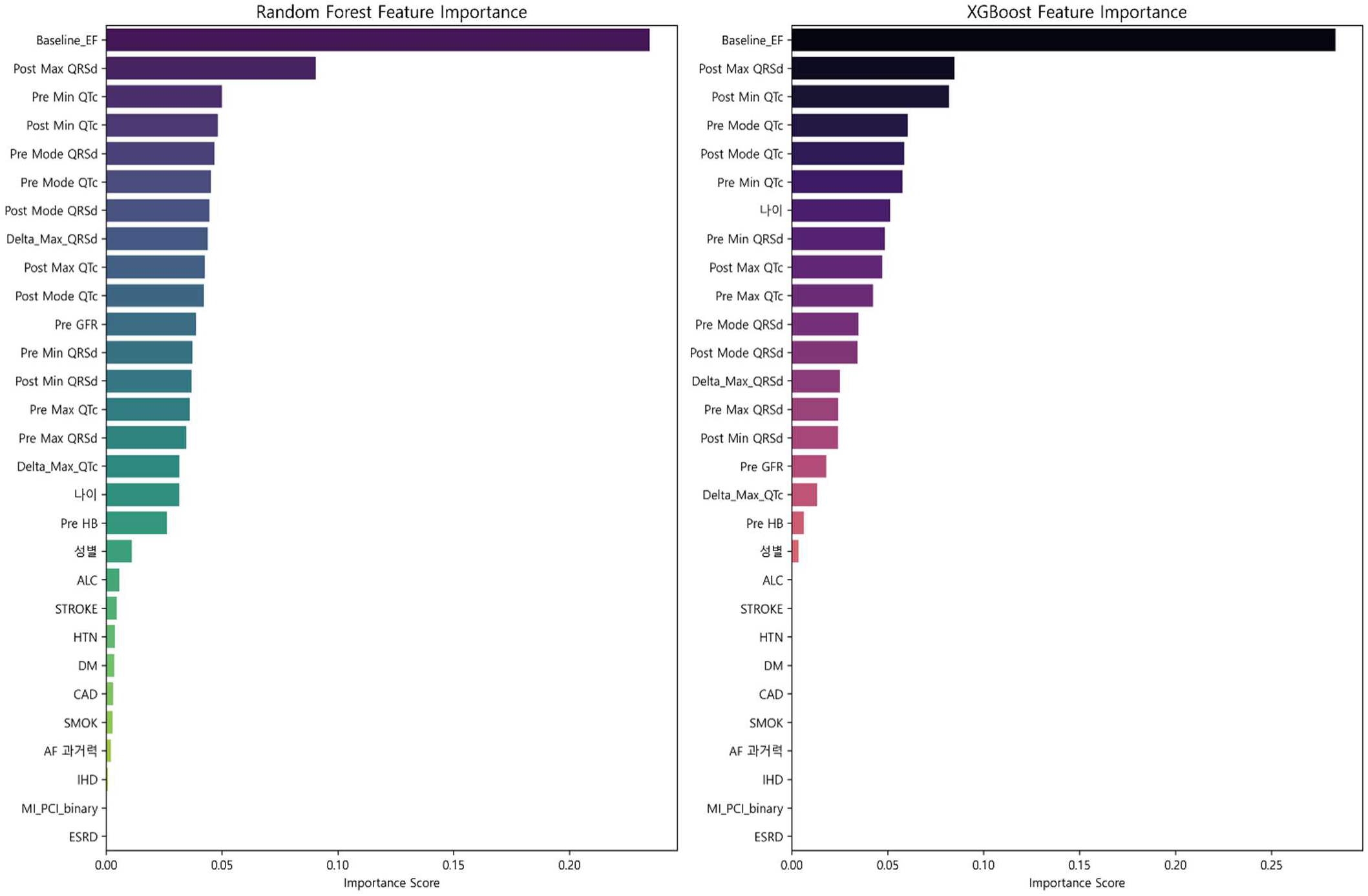
Feature importance analysis of the random forest and XGBoost models. Horizontal bar plots illustrating the relative contribution of predictor variables within the random forest (left) and XGBoost (right) models. Baseline ejection fraction (Baseline_EF) emerged as the most influential predictor in both models, followed by post-implant maximum QRS duration (Post Max QRSd). EF = ejection fraction; QRSd = QRS duration; QTc = corrected QT interval.

Post-implant maximum QRSd represented the second most influential feature in both models (random forest, 9.0%; XGBoost, 8.5%). Additional influential variables included post-implant minimum QTc interval (random forest, 4.8%; XGBoost, 8.2%), pre-implant mode QTc interval (random forest, 4.5%; XGBoost, 6.0%), and post-implant mode QTc interval (random forest, 4.2%; XGBoost, 5.9%).

### Risk Stratification and Survival Analysis

To evaluate the clinical utility of model-derived risk estimates, patients were stratified into high-risk and low-risk groups according to predicted probabilities generated by the best-performing model, XGBoost. As shown in Figure 3, the high-risk group demonstrated significantly lower PICM-free survival probabilities throughout follow-up compared with the low-risk group (log-rank *P* < 0.001). The Kaplan–Meier curves diverged early after pacemaker implantation and remained consistently separated during longitudinal follow-up, with the high-risk group exhibiting a substantially steeper decline in PICM-free survival.

**Figure 3.**
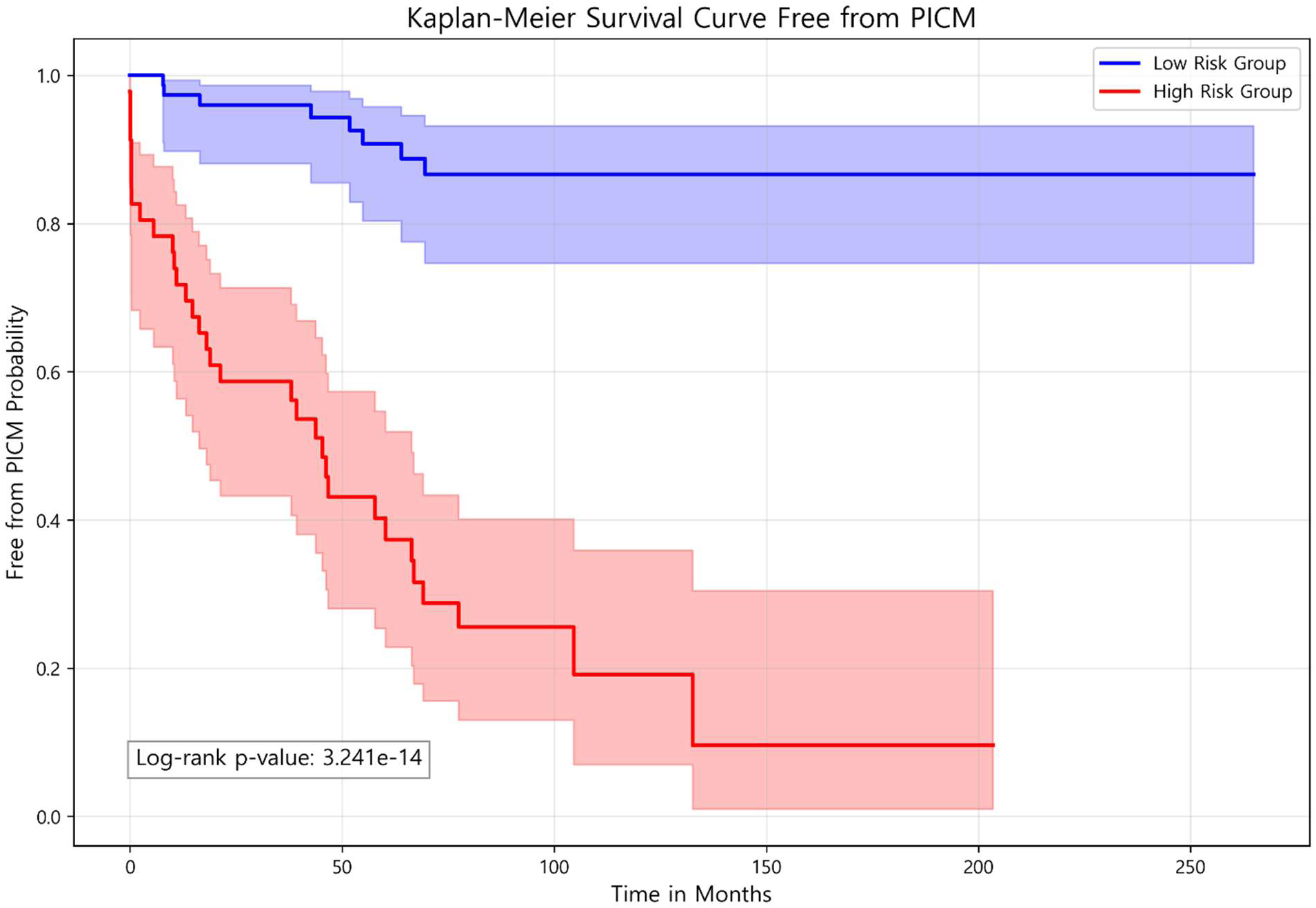
Kaplan–Meier curves for PICM-free survival stratified according to machine learning-derived risk groups. Patients were classified into high-risk (red) and low-risk (blue) groups based on predicted probabilities generated by the XGBoost classifier. The optimal classification threshold was determined by maximizing Youden’s J statistic. Shaded regions represent 95% confidence intervals. The high-risk group demonstrated significantly lower PICM-free survival than the low-risk group (log-rank P = 3.24 × 10−14). PICM = pacing-induced cardiomyopathy.

## Discussion

The present study demonstrates that ML-based models can effectively predict PICM occurrence in patients with AVB and underlying HFmrEF or HFpEF. This is the full-length, prospectively followed multicenter analysis to validate the predictive utility of ML approaches for estimating PICM risk in this real-world multicenter setting. These findings provide a clinically relevant and data-driven framework for individualized pacing strategy selection in patients with AVB requiring chronic RVP.

As discussed previously, CSP has emerged as a physiologically favorable alternative to conventional RVP; however, supporting evidence remains limited in pacing-dependent AVB with HFmrEF or HFpEF ^2,8,17,22^. In addition, the procedural complexity of CSP, including prolonged implantation times and higher lead revision rates, continues to limit its widespread adoption in routine clinical practice ^21,23^. Consequently, conventional RVP remains an important and frequently preferred pacing strategy for AVB patients considered to be at relatively low risk for PICM development. Accurate prediction of PICM risk is therefore essential to facilitate individualized selection between CSP and conventional RVP in AVB patients with HFmrEF or HFpEF.

Among the 125 patients included in the present analysis, 32.8% developed PICM during long-term follow-up. Although this incidence exceeds the pooled prevalence reported in previous meta-analyses, which approximates 12%, the discrepancy likely reflects differences in diagnostic definitions, the pacing-dependent rhythm of the present cohort (RVP burden ≥80%), and the prolonged follow-up duration of the current study ^4,24,25^.

The present findings demonstrated that male sex, pre-implant minimum QTc interval, and post-implant maximum QRSd were significantly associated with PICM occurrence, findings that are broadly consistent with prior investigations ^16,17,25,26^. The concordance of these findings with previous literature supports the external validity of the present results and reinforces the potential generalizability of the developed ML models. Among the evaluated ML algorithms, XGBoost demonstrated the highest discriminative performance, followed closely by random forest, with both ensemble-based approaches outperforming conventional logistic regression and SVM. Furthermore, XGBoost-based risk stratification effectively differentiated high-risk and low-risk groups, as demonstrated by marked separation of the Kaplan–Meier PICM-free survival curves.

The present results differ from our interim abstract^27^ in three respects, each methodologically explicable. First, the cohort was restricted to pacing-dependent AVB patients with HFpEF or HFmrEF and reclassified under a stricter PICM definition (post-implant LVEF <50%), yielding 41 events among 125 patients versus 49 among 154, with comparable incidence (32.8% vs 31.8%). Second, pre-implant QRS duration was significantly associated with PICM in the interim analysis (non-PICM 113±28ms vs. PICM 133±26ms, p<0.001) but not in the present analysis (maximum QRS duration; non-PICM 120 ± 31ms vs. PICM 127 ± 30ms, p=0.197), likely reflecting the exclusion of patients with lower RVP burden and the use of multiple ECG QRS duration recordings (mode, maximum, minimum) per patient; concurrently, pre-implant minimum QTc emerged as significant (421 ± 46ms vs. 446 ± 52ms, p=0.008). Third, intra-individual QRS duration variability, defined as the difference between the maximum and minimum QRS duration and evaluated as the principal exposure in our interim analysis, was retained as a prespecified candidate feature. Neither pre-implant nor post-implant QRS duration variability met the univariate selection threshold and neither was retained in the final models.

The present study has several important strengths. First, this full-length, prospective multicenter analysis provides a realistic estimate of achievable discrimination during long-term follow-up. Compared with previous deep learning approaches using CNN-based architectures for similar datasets, the ensemble-based methods used in the present study, particularly XGBoost and random forest, were more appropriately suited for structured tabular clinical variables, demonstrated substantially higher sensitivity (80.5% vs. 55.6%), and were developed within a more rigorous prospective multicenter framework.

Second, the incorporation of multiple ECG recordings per patient, including mode, maximum, and minimum QRSd, enabled more comprehensive characterization of each patient’s ECG profile than the single-ECG approach adopted in most prior investigations. This multi-recording ECG strategy may have enhanced the physiologic signal captured by the ML models and thereby contributed to the improved discriminative performance observed in the present analysis.

Third, feature importance analyses yielded clinically interpretable findings with strong pathophysiologic relevance. Baseline pre-implant LVEF emerged as the most influential variable in both the random forest (23.5%) and XGBoost (28.3%) models, supporting the concept that pre-existing functional remodeling of myocardium represents a major determinant of susceptibility to PICM development.

Post-implant maximum QRSd emerged as the second most influential feature, consistent with the established pathophysiologic principle that prolonged paced QRS complexes reflect greater ventricular dyssynchrony and consequently increased susceptibility to adverse electrical remodeling ^3,18^. Notably, although conventional Cox regression analysis did not identify baseline LVEF as a statistically significant independent variable, the ML-based models successfully captured its prognostic relevance within a multidimensional predictive framework. This discrepancy highlights a key advantage of ML approaches, namely their ability to identify non-linear associations and complex interactions among variables that may not be adequately captured through conventional regression-based analyses of isolated risk factors.

Despite these strengths, several limitations should be acknowledged. First, the relatively modest sample size may have affected the stability of model performance estimates. Although stratified four-fold cross-validation was implemented to reduce the risk of overfitting, external validation in larger independent cohorts remains necessary to establish broader model generalizability. Second, although the multicenter design enhanced external validity, interinstitutional variability in clinical practice patterns, echocardiographic measurement protocols, and multiple-ECG acquisition standards may have introduced heterogeneity and measurement noise into the dataset.

Third, incorporation of post-implant ECG parameters limits the utility of the current model as a purely pre-procedural risk stratification tool. Fourth, although robust cross-validation procedures were used to evaluate model performance, initial feature selection based on univariate statistical significance (*P* < 0.05) was performed before cross-validation using the full cohort.

Although this approach is consistent with conventional clinical statistical workflows for identifying clinically relevant variables, it may introduce a degree of data leakage within the ML framework and could potentially contribute to modest overestimation of predictive performance.

Given the established prognostic relevance of post-implant QRSd, one practical future strategy may involve obtaining QRSd measurements during temporary pacing before planned permanent pacemaker implantation in AVB patients with HFmrEF or HFpEF. Future prospective investigations could incorporate these pre-procedural ECG parameters into ML-based risk stratification frameworks and subsequently randomize eligible AVB patients with HFmrEF or HFpEF planned to CSP or conventional RVP. Such studies may provide the robust evidence necessary to optimize pacing modality selection and reduce the incidence of PICM.

## Conclusion

In patients with AVB with HFmrEF or HFpEF, ML-based models, particularly XGBoost, demonstrated clinically meaningful, internally validated discriminative performance relative to conventional approaches and effectively stratified patients according to clinically meaningful PICM risk. These findings support the potential clinical utility of ML-driven risk stratification to facilitate individualized chronic pacing strategy selection.

## Data Availability

The data that support the findings are available from the corresponding author upon reasonable request.

## Abbreviations

RVP: right ventricular pacing
PICM: pacing-induced cardiomyopathy
AVB: atrioventricular block
CSP: conduction system pacing
BiVP: biventricular pacing
CNN: convolutional neural network
ML: machine learning
AUROC: area under the receiver operating characteristic curve
SVM: support vector machine
XGBoost: extreme gradient boosting.

## Author Contribution

JW Lee: Data Curation, Formal Analysis, Project administration, Resource and Visualization U Do, HM Kim and SY Shin: Investigation and Methodology

MJ Choi and S Park: Software and Visualization

KW Kang: Conceptualization, Funding acquisition, supervision, validation, writing original draft, review and editing

## References

1. Akerström F, Pachón M, Puchol A, Jiménez-López J, Segovia D, Rodríguez-Padial L, Arias MA. Chronic right ventricular apical pacing: adverse effects and current therapeutic strategies to minimize them. Int J Cardiol. 2014;173:351–360. doi: 10.1016/j.ijcard.2014.03.079

2. Kusumoto FM, Schoenfeld MH, Barrett C, Edgerton JR, Ellenbogen KA, Gold MR, Goldschlager NF, Hamilton RM, Joglar JA, Kim RJ, et al. 2018 ACC/AHA/HRS Guideline on the Evaluation and Management of Patients With Bradycardia and Cardiac Conduction Delay: Executive Summary: A Report of the American College of Cardiology/American Heart Association Task Force on Clinical Practice Guidelines, and the Heart Rhythm Society. J Am Coll Cardiol. 2019;74:932–987. doi: 10.1016/j.jacc.2018.10.043

3. Ponnusamy SS, Syed T, Vijayaraman P. Pacing induced cardiomyopathy: recognition and management. Heart. 2023;109:1407–1415. doi: 10.1136/heartjnl-2022-321723

4. Somma V, Ha FJ, Palmer S, Mohamed U, Agarwal S. Pacing-induced cardiomyopathy: A systematic review and meta-analysis of definition, prevalence, risk factors, and management. Heart Rhythm. 2023;20:282–290. doi: 10.1016/j.hrthm.2022.09.019

5. Ferrick AM, Raj SR, Deneke T, Kojodjojo P, Lopez-Cabanillas N, Abe H, Boveda S, Chew DS, Choi JI, Dagres N, et al. 2023 HRS/EHRA/APHRS/LAHRS expert consensus statement on practical management of the remote device clinic. Heart Rhythm. 2023;20:e92–e144. doi: 10.1016/j.hrthm.2023.03.1525

6. Vijayaraman P, Subzposh FA, Naperkowski A. Atrioventricular node ablation and His bundle pacing. Europace. 2017;19:iv10–iv16. doi: 10.1093/europace/eux263

7. Reddy RK, Leong A, Naraen A, Liang Y, Ravendren A, Aggour H, Song Z, Saleh K, Kuramitsu S, Ahmad Y, et al. Conduction system versus right ventricular pacing for atrioventricular block: a reconstructed individual participant data meta-analysis. Eur Heart J. 2024;45. doi: 10.1093/eurheartj/ehae666.711

8. Vlachakis PK, Theofilis P, Apostolos A, Kordalis A, Sideris S, Leventopoulos G, Mountantonakis SE, Kollias G, Leontsinis I, Drakopoulou M, et al. Conduction system pacing in heart failure: Time for a paradigm shift? Heart Fail Rev. 2025;30:365–380. doi: 10.1007/s10741-024-10469-9

9. Glikson M, Nielsen JC, Kronborg MB, Michowitz Y, Auricchio A, Barbash IM, Barrabés JA, Boriani G, Braunschweig F, Brignole M, et al. 2021 ESC Guidelines on cardiac pacing and cardiac resynchronization therapy. Eur Heart J. 2021;42:3427–3520. doi: 10.1093/eurheartj/ehab364

10. Tóth AZ, Nagy L, Jenei C, Ráduly AP, Sándorfi G, Szabó KM, Kiss A, Nagy LT, Szilágyi GI, Csanádi Z. Conduction System Pacing Improved Cardiac Functions, Myocardial Work and Functional Capacity in Heart Failure with Reduced Ejection Fraction and Right Bundle Branch Block. J Clin Med. 2025;15. doi: 10.3390/jcm15010232

11. Glikson M, Burri H, Abdin A, Cano O, Curila K, De Pooter J, Diaz JC, Drossart I, Huang W, Israel CW, et al. European Society of Cardiology (ESC) clinical consensus statement on indications for conduction system pacing, with special contribution of the European Heart Rhythm Association of the ESC and endorsed by the Asia Pacific Heart Rhythm Society, the Canadian Heart Rhythm Society, the Heart Rhythm Society, and the Latin American Heart Rhythm Society. Europace. 2025;27. doi: 10.1093/europace/euaf050

12. Dickstein K, Vardas PE, Auricchio A, Daubert JC, Linde C, McMurray J, Ponikowski P, Priori SG, Sutton R, van Veldhuisen DJ. 2010 Focused Update of ESC Guidelines on device therapy in heart failure: an update of the 2008 ESC Guidelines for the diagnosis and treatment of acute and chronic heart failure and the 2007 ESC guidelines for cardiac and resynchronization therapy. Developed with the special contribution of the Heart Failure Association and the European Heart Rhythm Association. Eur Heart J. 2010;31:2677–2687. doi: 10.1093/eurheartj/ehq337

13. Huizar JF, Kaszala K, Tan A, Koneru J, Mankad P, Kron J, Ellenbogen KA. Abnormal Conduction-Induced Cardiomyopathy: JACC Review Topic of the Week. J Am Coll Cardiol. 2023;81:1192–1200. doi: 10.1016/j.jacc.2023.01.040

14. Zimerman A, Dal Forno A, Rohde LE, Ternes CM, Alves FD, Damiani LP, Martinelli-Filho M, Costa R, Fagundes AA, Barbosa RM, et al. Conduction System vs Biventricular Pacing in Heart Failure: The PhysioSync-HF Randomized Clinical Trial. JAMA Cardiol. 2026;11:360–368. doi: 10.1001/jamacardio.2026.0101

15. Khurshid S, Epstein AE, Verdino RJ, Lin D, Goldberg LR, Marchlinski FE, Frankel DS. Incidence and predictors of right ventricular pacing-induced cardiomyopathy. Heart Rhythm. 2014;11:1619–1625. doi: 10.1016/j.hrthm.2014.05.040

16. Kim JH, Kang KW, Chin JY, Kim TS, Park JH, Choi YJ. Major determinant of the occurrence of pacing-induced cardiomyopathy in complete atrioventricular block: a multicentre, retrospective analysis over a 15-year period in South Korea. BMJ Open. 2018;8:e019048. doi: 10.1136/bmjopen-2017-019048

17. Kiehl EL, Makki T, Kumar R, Gumber D, Kwon DH, Rickard JW, Kanj M, Wazni OM, Saliba WI, Varma N, et al. Incidence and predictors of right ventricular pacing-induced cardiomyopathy in patients with complete atrioventricular block and preserved left ventricular systolic function. Heart Rhythm. 2016;13:2272–2278. doi: 10.1016/j.hrthm.2016.09.027

18. Oida M, Mizutani T, Hasumi E, Fujiu K, Goto K, Kani K, Oshima T, Matsubara TJ, Shimizu Y, Oguri G, et al. Prediction of pacemaker-induced cardiomyopathy using a convolutional neural network based on clinical findings prior to pacemaker implantation. Sci Rep. 2024;14:6916. doi: 10.1038/s41598-024-57418-y

19. Kittleson MM, Panjrath GS, Amancherla K, Davis LL, Deswal A, Dixon DL, Januzzi JL, Jr., Yancy CW. 2023 ACC Expert Consensus Decision Pathway on Management of Heart Failure With Preserved Ejection Fraction: A Report of the American College of Cardiology Solution Set Oversight Committee. J Am Coll Cardiol. 2023;81:1835–1878. doi: 10.1016/j.jacc.2023.03.393

20. Bozkurt B, Coats AJ, Tsutsui H, Abdelhamid M, Adamopoulos S, Albert N, Anker SD, Atherton J, Bohm M, Butler J, et al. Universal Definition and Classification of Heart Failure: A Report of the Heart Failure Society of America, Heart Failure Association of the European Society of Cardiology, Japanese Heart Failure Society and Writing Committee of the Universal Definition of Heart Failure. J Card Fail. 2021;27:387–413. doi: 10.1016/j.cardfail.2021.01.022

21. Merchant FM, Mittal S. Pacing induced cardiomyopathy. J Cardiovasc Electrophysiol. 2020;31:286–292. doi: 10.1111/jce.14277

22. Abdin A, Aktaa S, Vukadinović D, Arbelo E, Burri H, Glikson M, Meyer C, Munyombwe T, Nielsen JC, Ukena C, et al. Outcomes of conduction system pacing compared to right ventricular pacing as a primary strategy for treating bradyarrhythmia: systematic review and meta-analysis. Clin Res Cardiol. 2022;111:1198–1209. doi: 10.1007/s00392-021-01927-7

23. Michowitz Y, Glikson M, Burri H. The ‘10 Commandments’ from the 2025 ESC/EHRA Clinical Consensus Statement on Indications for Conduction System Pacing (CSP). Eur Heart J. 2025;46:2844–2846. doi: 10.1093/eurheartj/ehaf371

24. Boriani G, Vitolo M, Proietti M. Cardiomyopathy associated with long-term right ventricular pacing: an intriguing clinical issue. Eur J Heart Fail. 2019;21:652–654. doi: 10.1002/ejhf.1449

25. Dreger H, Maethner K, Bondke H, Baumann G, Melzer C. Pacing-induced cardiomyopathy in patients with right ventricular stimulation for >15 years. Europace. 2012;14:238–242. doi: 10.1093/europace/eur258

26. Khurshid S, Liang JJ, Owens A, Lin D, Schaller R, Epstein AE, Marchlinski FE, Frankel DS. Longer Paced QRS Duration is Associated With Increased Prevalence of Right Ventricular Pacing-Induced Cardiomyopathy. J Cardiovasc Electrophysiol. 2016;27:1174–1179. doi: 10.1111/jce.13045

27. Lee JW, Do U, Shin S, Park S, Kang KW. Artificial intelligence-assisted prediction for occurrence of PICM in the AV block. J Card Fail. 2026;32(1):197 (abstract 051)

